# Central role of glycosylation processes in human genetic susceptibility to SARS-CoV-2 infections with Omicron variants

**DOI:** 10.1101/2024.11.21.24317689

**Authors:** Frank Geller, Xiaoping Wu, Vilma Lammi, Erik Abner, Jesse Tyler Valliere, Katerina Nastou, Morten Rasmussen, Niklas Worm Andersson, Liam Quinn, DBDS Genomic Consortium, Bitten Aagaard, Karina Banasik, Sofie Bliddal, Lasse Boding, Søren Brunak, Nanna Brøns, Jonas Bybjerg-Grauholm, Lea Arregui Nordahl Christoffersen, Maria Didriksen, Khoa Manh Dinh, Christian Erikstrup, Ulla Feldt-Rasmussen, Kirsten Grønbæk, Kathrine Agergård Kaspersen, Christina Mikkelsen, Claus Henrik Nielsen, Henriette Svarre Nielsen, Susanne Dam Nielsen, Janna Nissen, Celia Burgos Sequeros, Niels Tommerup, Henrik Ullum, Estonian Biobank Research Team, FinnGen, Lampros Spiliopoulos, Peter Bager, Anders Hviid, Erik Sørensen, Ole Birger Pedersen, Jacqueline M Lane, Ria Lassaunière, Hanna M Ollila, Sisse Rye Ostrowski, Bjarke Feenstra

## Abstract

The host genetics of SARS-CoV-2 has previously been studied based on cases from the earlier waves of the pandemic in 2020 and 2021, identifying 51 genomic loci associated with infection and/or severity. SARS-CoV-2 has shown rapid sequence evolution increasing transmissibility, particularly for Omicron variants, which raises the question whether this affected the host genetic factors. We performed a genome-wide association study of SARS-CoV-2 infection with Omicron variants including more than 150,000 cases from four cohorts. We identified 13 genome-wide significant loci, of which only five were previously described as associated with SARS-CoV-2 infection. The strongest signal was a single nucleotide polymorphism (SNP) intronic of *ST6GAL1*, a gene affecting immune development and function, and connected to three other associated loci (harboring *MUC1, MUC5AC* and *MUC16*) through O-glycan biosynthesis. We also found further evidence for an involvement of blood group systems in SARS-CoV-2 infection, as we observed association 1) for a different lead SNP in the *ABO* locus indicating a protective effect of blood group B against Omicron infection, 2) for the *FUT2* SNP tagging secretor status also reported for SARS-CoV-2 infection with earlier variants, and 3) for the strongest expression quantitative trait locus (eQTL) for *FUT3* (Lewis gene). Our study provides robust evidence for individual genetic variation related to glycosylation translating into susceptibility to SARS-CoV-2 infections with Omicron variants.

## Introduction

According to numbers from the World Health Organization, severe acute respiratory syndrome coronavirus 2 (SARS-CoV-2) has by now caused more than 770 million cases of COVID-19, resulting in more than 7 million deaths^1^. The largest genetic study on susceptibility to SARS-CoV-2 infection was a genome-wide association study (GWAS) by the COVID-19 Host Genetics Initiative (HGI), meta-analyzing up to 219,692 cases and over 3 million controls, which identified 51 genetic loci^2^ associated with infection and/or two other outcomes related to COVID-19 disease severity. However, that study was built on a data freeze from December 2021 just after the detection of Omicron in November 2021, thus only including infections with earlier (pre-Omicron) SARS-CoV-2 variants. The evolution of the virus gave rise to multiple mutations that affected, amongst others, the transmissibility of the virus^3^. Omicron variants showed more mutations than earlier variants and, within a few months, infected far more individuals worldwide than all the earlier variants combined.

Given these substantial changes observed for the virus, we decided to investigate the corresponding host genetics by performing a GWAS of SARS-CoV-2 infection with Omicron variants in more than 150,000 cases and more than 500,000 controls without a known SARS-CoV-2 infection by combining data from four cohorts in a meta-analysis.

## Results

### GWAS of Omicron infection vs. no infection

In our main analysis, we compared SARS-CoV-2 infection with Omicron variants (proxied by the first reported infection observed in a period where Omicron variants were dominating in the study cohorts, that was after the start of 2022) vs. controls with no known SARS-CoV-2 infection from electronic health records, viral testing or questionnaire data in the covered time period (see **Online Methods** for further details). To simplify matters, genetic variants are denoted as SNPs throughout the manuscript, so that the term variant always refers to variation in SARS-CoV-2.

We performed a meta-analysis of four GWAS with a total of 151,825 cases and 556,568 controls (see **Figure 1** for Manhattan plot) and identified 13 genome-wide significant loci, of which 8 represent novel associations for SARS-CoV-2 infection (**Table 1**). Four of the corresponding lead SNPs had proxies among the previously reported SNPs associated with SARS-CoV-2 infection related to earlier variants (r^2^ > 0.6), and for the *SLC6A20* locus, the lead SNP reported for the earlier variants was in the 95% credible set of our GWAS signal (rs73062389, *P* = 8.9 × 10^−33^ in our study). Two of these loci had been assigned to the pathway “entry defense in airway mucus” (nearby genes *MUC1* and *MUC16*) and one to “viral entry and innate immunity” (*SLC6A20*)^2^. The other two loci previously reported in the context of earlier variants identified in our meta-analysis were represented by rs13100262 (*RPL24*) and rs492602 (*FUT2*). The protective allele rs492602-G is related to non-secretor status, which confers resistance to certain viral infections (e.g. Norovirus, Rotavirus) and susceptibility to other conditions (e.g. mumps, measles, kidney disease)^4^.

**Table 1.**
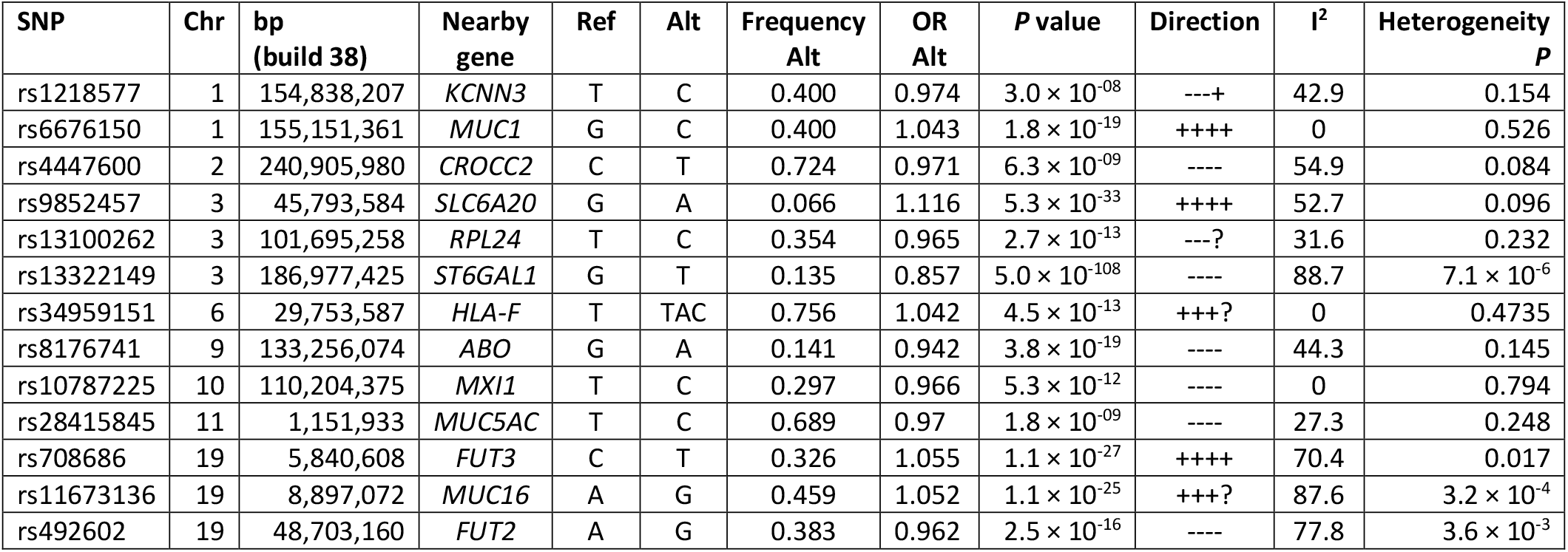
Associated loci from the meta-analysis. The order of the studies in the direction column is according to effective sample size FinnGen, Estonian Biobank, Denmark and Mass General Brigham Biobank.

**Figure 1:**
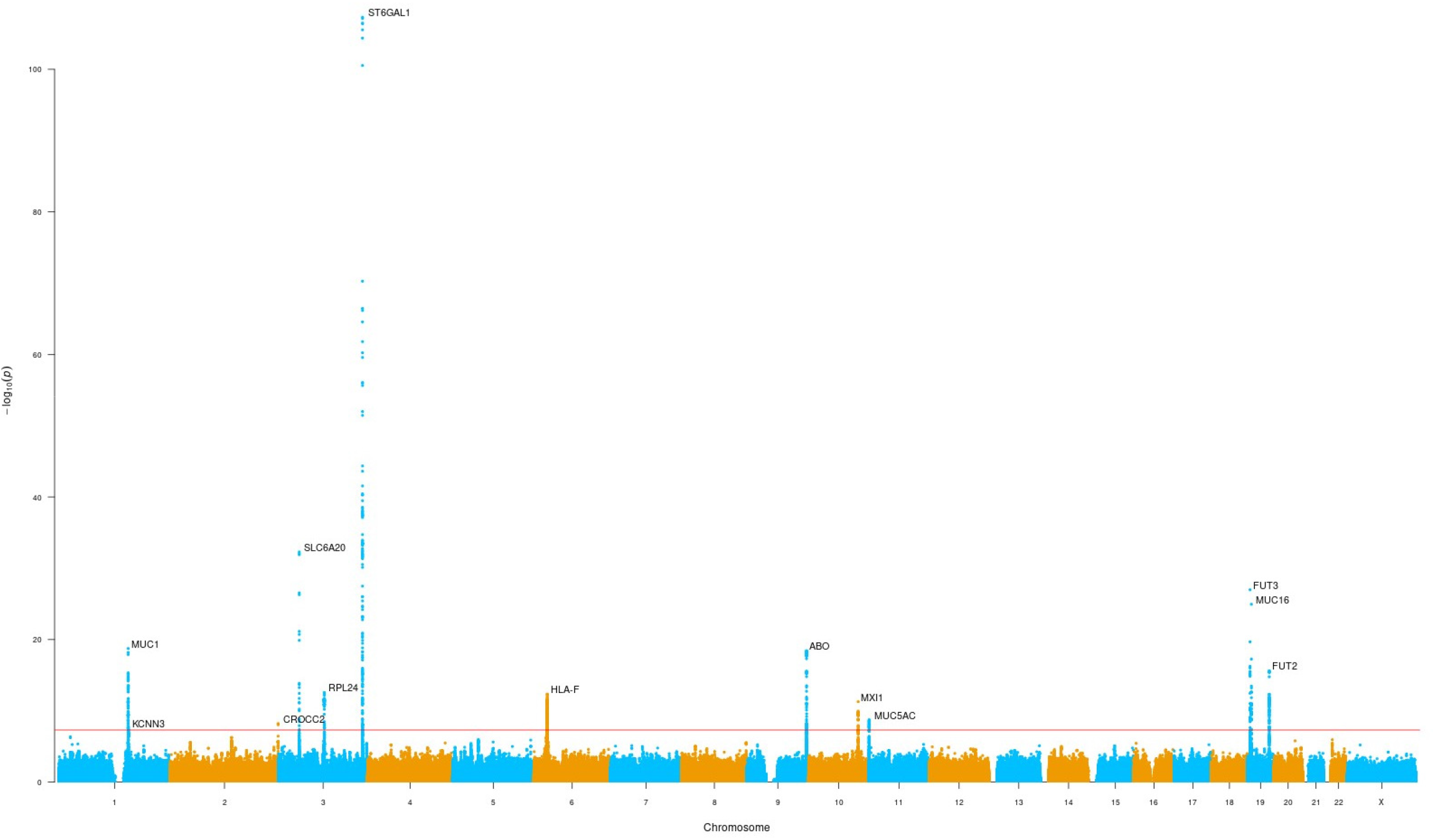
Manhattan plot for GWAS of Omicron infection vs. no known infection.

The most significant finding was the intronic SNP rs13322149 (odds ratio (OR) for minor allele T: 0.857, *P* = 5 × 10^−108^) in *ST6GAL1* (ST6 beta-galactoside alpha-2,6-sialyltransferase 1), a gene affecting immune development and function^5^. The encoded protein adds terminal α 2,6-sialic acids to galactose-containing N-linked glycans. A recent multi-ancestry GWAS of influenza infection also identified a protective effect for the minor allele T^6^. The strong association with influenza was further seen in phenome-wide association (PheWAS) results from the most recent FinnGen cohort (FinnGen release 12 (https://www.finngen.fi/en), with an OR of 0.889 for rs13322149-T (*P* = 5.2 × 10^−10^, 11,558 cases vs. 415,538 controls, r^2^ = 0.965 with the FinnGen influenza lead SNP rs55958900). The second new locus was represented by rs708686 (OR for allele T: 1.055, *P* = 1.1 × 10^−27^), located intergenic between the fucosyltransferases *FUT6* and *FUT3* (Lewis gene) and from the same gene family as *FUT2* harboring rs492602 mentioned above. In FinnGen release 12, the risk allele for Omicron infection rs708686-T was reported as lead SNP in cholelithiasis (OR = 1.103, *P* = 9.6 × 10^−41^, 49,834 cases vs. 437,418 controls), as well as in viral and other specified intestinal infections (OR = 0.913, *P* = 4.4 × 10^−10^, 11,050 cases vs. 444,292 controls), and it was the strongest eQTL for FUT3 levels (*β* = -0.657, *P* = 3 × 10^−126^) in a proteomics study^7^. The third SNP rs10787225 (OR for C: 0.966, *P* = 5.3 × 10^−12^), is located about 3 kilobase (kb) upstream of *MXI1* (MAX interactor 1), a region with GWAS findings for, amongst others, blood pressure^8^ and blood cell phenotypes^9^, but the previously identified SNPs are not in linkage disequilibrium (LD) with our lead SNP. Additional novel associations include rs4447600 (OR for T: 0.971, *P* = 6.3 × 10^−9^) on 2q37.3, which is in LD with rs6437219 (r^2^ = 0.64 in the Danish study population), associated with forced vital capacity (FVC)^10^. Reduced FVC can indicate reduced lung function, and at this locus the allele linked to reduced FVC is in phase with the allele conferring an increased risk of Omicron infection. The genetic association at the *ABO* locus changed drastically, as the previously reported SNP rs505922 linked to a protective effect of blood group O for earlier variants^2^ has changed direction of effect and no longer showed the strongest association (OR for major allele T 1.022, *P* = 4.8 × 10^−6^). Instead, rs8176741 (OR for minor allele A: 0.942, *P* = 3.8 × 10^−19^, r^2^ = 0.159 with rs505922 in European ancestry) was the lead SNP and as it tags blood group B, a protective effect of blood group B against SARS-CoV-2 infection with Omicron variants can be inferred.

The human leukocyte antigen (HLA) region and the *MUC5AC* locus have previously shown association with COVID-19-severity^2^, but with SNPs that show no strong LD to the lead SNP in this GWAS (r^2^ < 0.3). Our top HLA SNP rs34959151 (OR for TAC: 1.042, *P* = 4.5 × 10^−13^) is in strong LD with rs1736924 (r^2^ = 0.989 in the Danish study population), which tags HLA-F*01:03^11^, and there is growing evidence that HLA-F plays a significant role in immune modulation and viral infection^12^.

Our finding near *MUC5AC* (rs28415845, OR for C: 0.97, *P* = 1.8 × 10^−9^) adds further evidence for the role of mucins in protecting against infection with Omicron variants^13^. Finally, rs1218577 (OR for C: 0.974, *P* = 3 × 10^−8^) is located near *KCNN3*, not too far from the *MUC1* locus. However, the SNP is located more than 300 kb apart from rs6676150 in a different LD block (D’ = 0.162, r^2^ = 0.0096) and deserves further attention. Four lead SNPs showed signs of heterogeneity of effect between the study groups with *P* < 0.05 in Cochran’s Q-test and I^2^ > 60. However, all four SNPs have *P* values well below the genome-wide significance threshold and the heterogeneity is mainly a result of substantially stronger effect estimates in the Danish cohort (see **Supplementary Figure 1** for forest plots). This is probably a consequence of Denmark being one of the countries which had extremely high test activity with easily accessible testing for the whole population^14^; all cases in the study group were identified by positive PCR-test and controls were selected based on a negative PCR-test and a test history without any positive test.

### Relation to GWAS of earlier SARS-CoV-2 variants

We looked up all 51 SNPs reported by the HGI as associated with SARS-CoV-2 infection and/or hospitalization in their Supplementary Table 5^2^ (**Supplementary Table 1**), where we observed a comparable effect for rs190509934 close to *ACE2*, with *P* = 8.9 × 10^−7^ in the FinnGen cohort, indicating that this relatively rare SNP did not reach genome-wide significance in our study due to the reduced power from being reported in one cohort only.

To overcome the problems inherent to comparing two GWAS meta-analyses on different phenotypes with different cohorts involved, we investigated differences to the genetic findings for earlier variants by performing a second GWAS in our cohorts. Again, we used cases of SARS-CoV-2 infection with Omicron variants, but now vs. controls with a SARS-CoV-2 infection before Omicron variants had notable case numbers (“earlier variants”, i.e. infection before December 2021, N = 87,212). The results we obtained for the lead SNPs from **Table 1** (**Supplementary Table 2**) underlined the emergence of the *ST6GAL1* locus (*P* = 2 × 10^−49^), and the new lead SNP at the *ABO* locus (*P* = 1.6 × 10^−18^). The difference for the previously reported *ABO* SNP rs505922 was even stronger (*P* = 1.7 × 10^−30^), confirming the protective effect observed in earlier variants. For the other lead SNPs, *P* ranged from 9.4 × 10^−7^ to 0.82, with the most significant difference caused by a stronger effect related to Omicron variants at the previously reported *MUC16* locus.

### Relation to GWAS of breakthrough infections

A recent GWAS of SARS-CoV-2 breakthrough infections in the UK Biobank identified 10 loci^15^, of which 8 overlap with our findings (**Supplementary Table 3**), including all five loci that also were in common with the GWAS of infection with earlier SARS-CoV-2 variants. Among the remaining five loci associated with Omicron infection in our study, lead SNPs at four loci had P < 0.001 in the GWAS of breakthrough infections, only for the secondary signal at the chromosome 1 locus there was no sign of association. The lead SNPs at the two remaining loci in the GWAS of breakthrough infections had attenuated effect sizes and only reached nominal significance in our meta-analysis. The UK biobank study did not specify the time period where the breakthrough infections occurred, but given the overall large fraction of Omicron infections among all SARS-CoV-2 breakthrough infections, it can be expected that Omicron accounted for the majority of cases.

### Relation to GWAS of influenza

We looked up our genome-wide significant loci in a recent GWAS of influenza (**Supplementary Table 4)**, a study that also reported rs13322149 near *ST6GAL1* as lead SNP with a similar effect (OR for T 0.888, *P* = 3.6 × 10^−19^) ^6^.

In a total of 14 comparisons (including the only other lead SNP rs2837113 from the influenza GWAS), we observed two more of our loci reaching the adjusted significance level of 4.2 × 10^−3^ for influenza: rs6676150 (OR for C 1.038, *P* = 1.1 × 10^−6^) and the proxy SNP rs73005873 (OR for C 1.033, *P* = 5.0 × 10^−5^) near *MUC1* and *MUC16*, respectively, with consistent directions of effects between the studies. Contrary, the second lead SNP identified in the influenza GWAS (rs2837113, *B3GALT5* locus, OR for A 0.915, *P* = 4.1 × 10^−32^) went the opposite direction for Omicron (OR for A 1.016, *P* = 7.5 × 10^−4^).

### Gene-set and pathway analysis

We followed up on our GWAS with FUMA (v1.5.2)^16^ for a comprehensive integration of our results with public resources, including functional annotation, eQTL and chromatin interaction mapping, as well as additional gene-based, pathway and tissue enrichment tests (full results available online). On top of the entries from the GWAS catalogue for SNPs in LD with the lead SNPs provided by FUMA, we performed a comprehensive phenome-wide association study in 2,470 phenotypes provided in FinnGen release 12 for the lead SNPs (**Supplementary Table 5**), where the posterior inclusion probability (PIP) calculated with SuSie^17^ gives an indication whether our lead SNP is causal for the observed phenotype association. The MAGMA (v1.08)^18^ gene-set analysis identified the Reactome set “Termination of O-glycan biosynthesis” as the top set among a variety of 17,012 gene-sets (*P* = 6.8 × 10^−7^). Among the 23 genes in this gene-set are *ST6GAL1* and several mucin genes, including *MUC1, MUC5AC*, and *MUC16*, located in three distinct genome-wide significant loci in our study. Looking at the specific analysis for Reactome gene-sets in the secondary analysis GENE2FUNC, 10 gene-sets had an adjusted *P* < 0.05, eight of which are related to mucins/glycosylation (**Supplementary Table 6**).

### Functional protein association network analysis

We selected one gene for each of our 13 GWAS loci, and obtained functional protein association networks from STRING database v12^19^ for the corresponding proteins. Seven of the 13 proteins had functional associations above the default medium confidence score threshold of 0.4, and MUC1, MUC16 and MUC5AC also interacted physically on top of their functional associations (**Figure 2a**). As mentioned above, ST6GAL1 and the three mucins are all involved in the Reactome^20^ pathway “Termination of O-glycan biosynthesis”, where ST6GAL1 transfers sialic acid to galactose-containing acceptor substrates (here the mucins), and the connections were mainly a result of their involvement in this pathway. The connected component in this network also included FUT2, FUT3 and ABO, with the significant functional enrichment resulting from their involvement in the KEGG^21^ pathway “Glycosphingolipid biosynthesis – lacto and neolacto series” (the only significant pathway in the specific analysis for KEGG gene-sets in the secondary MAGMA analysis GENE2FUNC, adjusted *P* = 2.2 × 10^−4^). On top of these well-established connections, there were some weaker associations between ST6GAL1, FUT2 and FUT3, as well as between FUT3 and MUC1. The former connections were due to these proteins regulating glycosylation processes^22,23^, while the association between FUT3 and MUC1 was observed in aberrant glycosylation processes^22^. We expanded the network with 15 additional interactors at the maximum selectivity value of 1 to focus on proteins that primarily interact with the current network. For four of the identified interactors the corresponding gene was in a genomic locus already covered. The resulting highly specific network (**Figure 2b**) showed that the expansion added more proteins to the pathways already identified above. Amongst these proteins, another sialyltransferase (ST3GAL4) was involved in both pathways, and represents a strong link between the two sets of proteins.

**Figure 2:**
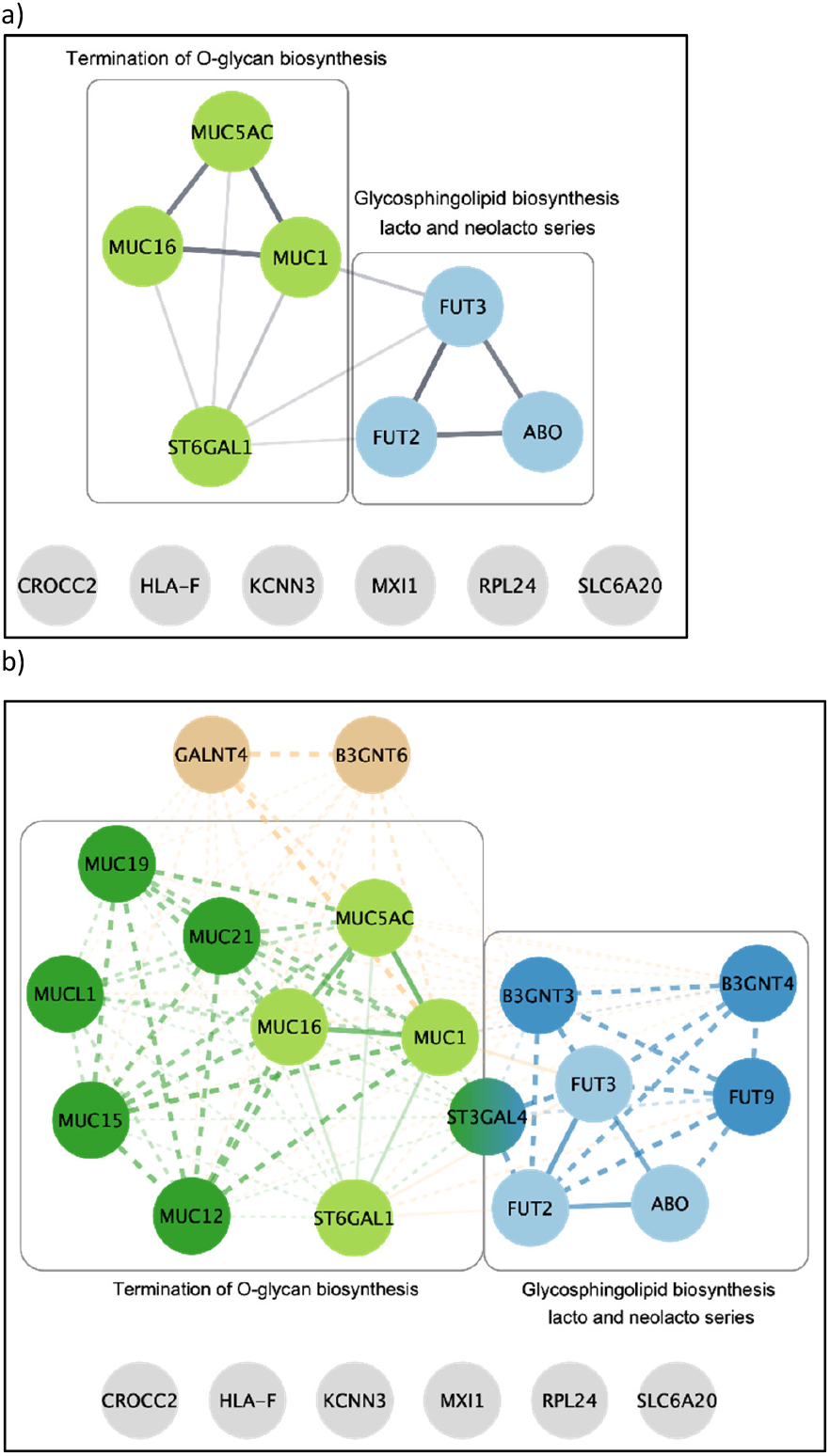
STRING networks. a) STRING network for 13 genes linked to the GWAS lead SNPs. Proteins involved in the Termination of O-glycan biosynthesis pathway are colored in light green, while proteins involved in Glycosphingolipid biosynthesis – lacto and neolacto series are colored in light blue. The two sets of proteins form a connected component with ST6GAL1 and FUT3 acting as the main bridges. The edge width is indicative of the confidence score for each association, with thicker edges denoting higher confidence scores. Physical protein associations are denoted with dotted lines. b) STRING network expanded with 15 additional interactors using a selectivity parameter of 1.0. Four interactors are removed because the corresponding genes were located in genomic loci already covered (FUT5, FUT6, MUC22, MUC3A). Additional proteins that belong to the Termination of O-glycan biosynthesis pathway are shown in dark green and additional proteins that belong to the Glycosphingolipid biosynthesis – lacto and neolacto series pathway are shown in dark blue. The addition of the extra proteins leads to a heavily interconnected network and for this reason we have selected a special coloring scheme to distinguish between the different edges in the network. Solid lines represent associations between the 13 original genes and dashed lines represent associations from the 11 additional genes. Green edges show associations between the genes involved in Termination of O-glycan biosynthesis pathway, blue edges show associations between the genes involved in Glycosphingolipid biosynthesis -lacto and neolacto series pathway and orange lines represent other associations.

### Heritability and genetic correlations

We estimated heritability from our GWAS at the liability scale assuming a prevalence of 0.5 as 0.024 (95%CI: 0.018–0.029), slightly higher than the heritability estimates for the HGI GWAS of infection vs. population controls in European ancestry (estimates for different scenarios were all below 0.019)^2^.

The genetic correlation between our GWAS for infection with Omicron variants and the publicly available meta-analysis results for infection with earlier variants from the HGI for individuals of European ancestry was estimated as r_g_ = 0.549 (95%CI: 0.342–0.757, *P* = 2.06 × 10^−7^). We also investigated genetic correlations of our GWAS with GWAS for 1,461 traits implemented in the Complex Traits Genetics Virtual Lab (CTG-VL, https://vl.genoma.io/), most results coming from the UK Biobank. With schizophrenia, r_g_ = -0.265 (95%CI: - 0.347– -0.182, *P* = 2.95 × 10^−10^), and asthma, r_g_ = 0.289 (95%CI: 0.187–0.39, *P* = 2.67 × 10^−8^), two serious health conditions were among the traits reaching the adjusted significance level of 3.4 × 10^−5^ (**Supplementary Table 7**).

## Discussion

We performed a GWAS of SARS-CoV-2 infection with Omicron variants in more than 150,000 cases and more than 500,000 controls without a known SARS-CoV-2 infection from four cohorts and identified 13 genome-wide significant loci. Our study investigated infection during the Omicron period in general, since information on the sub-variants of Omicron that regularly emerge was not available at an individual level. However, more than 70% of our cases were from the first six months of 2022, where BA variants were dominating in the study populations (see **Supplementary Figures 2 and 3**). It is notable that our findings are corroborated by a recent GWAS of breakthrough infections^15^, likely to be dominated by Omicron infections. Breakthrough and Omicron infections are closely related in large parts of Europe and the US, as the extensive vaccination programs rolled out in 2021 exerted strong selective pressure on the SARS-CoV-2 virus and were followed by the evolution and rapid spread of Omicron variants.

Among our findings, the most significant SNP is an intronic transversion mutation (rs13322149: G>T) located within the 148 kb β-galactoside α-2,6-sialyltransferase 1 (*ST6GAL1*) gene. ST6GAL1 catalyzes the addition of terminal α 2,6-sialic acids to galactose-containing N-linked glycans and is highly expressed in the liver, glandular cells in the prostate, collecting ducts and distal tubules in the kidneys, and germinal centers in lymph nodes (https://www.proteinatlas.org/ENSG00000073849-ST6GAL1/tissue). Expression of *ST6GAL1* also enhances the concentration of 6-linked sialic acid receptors accessible to influenza virus on the cell surface^24^. Based on knowledge from other corona viruses (including MERS-CoV recognizing α2,3 sialic acids and to a lesser extent the α2,6 sialic acids and sulfated sialyl-Lewis^X^ for binding preference), a role of O-acetylated sialic acids in the entry of SARS CoV-2 into the host cell has been postulated early in the pandemic^25^, resulting in multiple studies on the topic in a short time^26^.

It is evident from *in vitro* and *in vivo* studies that the emergence of Omicron changed the interaction of SARS-CoV-2 with the host. Compared to the ancestral B.1. lineage virus and the Delta variant, Omicron viral entry and infection is significantly attenuated in immortalized lung cell lines^27,28,29^ and human-derived lung organoids^30^, but increased in human-derived upper airway organoids^27^. In transgenic mice and Syrian hamsters, Omicron is also less pathogenic with reduced infection and pathology in the lower airways^31^, but with greater affinity for tracheal cells^32^. The mechanism underlying this tropism shift is not fully understood. Here, the association of our *ST6GAL1* SNP rs1334922 with reduced infection risk for Omicron, but not pre-Omicron variants, suggests an involvement of α2,6 sialic acids that emerged with the evolution of this SARS-CoV-2 variant. Considering that the same *ST6GAL1* lead SNP is protective against influenza infection, a virus that enters cells through binding α2,6 sialic acids, and the dependency of other beta coronaviruses on sialic acids for host cell entry (reviewed by Sun^26^) warrants a re-evaluation of the role of sialic acids in SARS-CoV-2 host cell entry for Omicron variants.

In addition to a role for host cell glycosylation in viral entry, the SARS-CoV-2 spike protein is itself heavily glycosylated, with 22 N-glycosylation sites per monomer. These glycans shield the protein from the host’s humoral immune response^33,34^ and are generally conserved across earlier and later variants, including Omicron^35,36^. However, Omicron has decreased sialylation of these glycans^35,37^, which is speculated to reduce electrostatic repulsion and steric hindrance when binding to the ACE2 receptor and ultimately promote stronger binding between the Omicron spike and this host receptor^38,39^. Glycosylation near the furin cleavage site can also regulate viral activity^40,41^, where sialic acid occupancy on O-glycans decreases furin activity by up to 65%^42^. Together, these results suggest that a reduction in sialic acid levels on the spike protein can enhance the infectivity of SARS-CoV-2 through improved binding to the ACE2 receptor and increased furin activity.

Gene-set analysis linked *ST6GAL1* to mucin genes, and our GWAS identified three loci with mucin candidate genes (*MUC1, MUC5AC*, and *MUC16*), showing that the biological pathway airway defense in mucus linked to infections with earlier SARS-CoV-2 variants^2^ also plays an important role in relation to Omicron variants. A recent GWAS of influenza identified two SNPs associated at genome-wide significance and, based on SARS-CoV-2 GWAS results for earlier variants, concluded that the genetic architectures of COVID-19 and influenza are mostly distinct. Our results provide nuance, as our *ST6GAL1* SNP for Omicron infection was one of the two lead SNPs for influenza infection and showed a similar effect. Additionally, two of our three mucin loci had suggestive findings in the influenza GWAS.

Additional evidence for a connection between blood group systems and SARS-CoV-2 infection was obtained by three associated loci, finding the same association at the *FUT2* locus determining secretor status as described for earlier variants, identifying a new locus near *FUT3*, and observing substantial differences at the ABO locus, where the lead SNP indicates a protective effect of blood group B. We want to stress that our results did not contradict the protective effect of blood group O reported for earlier variants, as the previously associated SNP was the one showing the largest difference between cases infected with Omicron variants vs. controls infected with earlier variants.

In conclusion, our study indicates that the human genetic architecture of SARS-CoV-2 infection is under constant development and updated GWAS analyses for periods where certain variants dominate can give further insights into the involved biological mechanisms. Our results indicate that processes related to glycosylation are particularly relevant for infections with Omicron variants. Experimental studies comparing the infectivity of different SARS-CoV-2 variants in relation to host cell expression of *ST6GAL1* and other mediators of glycosylation are needed to decipher the underlying biology.

## Methods

### Denmark

For the Danish cohort we combined genotype data from the Copenhagen Hospital Biobank and the Danish Blood Donor Study with information on SARS-CoV-2 infection from the EFTER-COVID study^43^. In short, the EFTER-COVID study invited individuals above 15 years of age with a RT-PCR-test for SARS-CoV-2 infection between 1^st^ September 2020 and 21^st^ February 2023 to fill in a baseline and several follow-up questionnaires. Cases for SARS-CoV-2 infection with Omicron variants had either a positive test after 28^th^ December 2021, when more than 90% of new infections were Omicron, or an Omicron infection confirmed by variant-specific PCR-test earlier in December 2021. Controls were individuals with a negative PCR-test related to the EFTER-COVID study and no positive test-result for any test in the database. For the comparison with earlier infections, controls either were defined as having a positive test before Omicron infections were observed in Denmark (21^st^ November 2021) or infection with a non-Omicron variant confirmed by variant-specific PCR-test in December 2021. Basic descriptive statistics on age and sex of cases and controls from all cohorts are given in **Supplementary Table 8**. Genetic data for the Copenhagen Hospital Biobank and the Danish Blood Donor Study were available from genotyping with Illumina Global Screening Arrays and subsequent imputation as previously described^44,45^. Data cleaning steps included filtering out individuals who were of non-European genetic ancestries (by removing outliers in a principal component analysis, deviating more than 5 standard deviations from one of the first 5 principal components), related (relatedness coefficient greater than 0.0883), having discordant sex information (chromosome aneuploidies or difference between reported sex and genetically inferred sex), were outliers for heterozygosity, or having more than 3% missing genotypes. Case-control GWAS analyses were performed with REGENIE (version 2.2.4)^46^ under an additive model, adjusting for sex and the first 10 principal components.

### Estonian Biobank

The Estonian Biobank (EstBB) is a population-based biobank with 212,955 participants in the current data freeze (2024v1). All biobank participants have signed a broad informed consent form and information on ICD-10 codes is obtained via regular linking with the national Health Insurance Fund and other relevant databases, with majority of the electronic health records (EHRs) having been collected since 2004^47^. COVID-19 data was acquired from EHRs (ICD-10 U07* category), with diagnoses between 1^st^ March 2020 till 30^th^ November 2021 being considered as cases with non-Omicron variants, while cases from 1^st^ January 2022 till 31^st^ December 2022 were considered as Omicron cases. Participants with diagnoses from both periods were excluded. Controls without any U07* category diagnoses were considered as healthy.

All EstBB participants have been genotyped at the Core Genotyping Lab of the Institute of Genomics, University of Tartu, using Illumina Global Screening Array v3.0_EST. Samples were genotyped and PLINK format files were created using Illumina GenomeStudio v2.0.4. Individuals were excluded from the analysis if their call-rate was < 95%, if they were outliers of the absolute value of heterozygosity (> 3SD from the mean) or if sex defined based on heterozygosity of X chromosome did not match sex in phenotype data^48^. Before imputation, variants were filtered by call-rate < 95%, HWE p-value < 1e-4 (autosomal variants only), and minor allele frequency < 1%. Genotyped variant positions were in build 37 and were lifted over to build 38 using Picard. Phasing was performed using the Beagle v5.4 software^49^. Imputation was performed with Beagle v5.4 software (beagle.22Jul22.46e.jar) and default settings. Dataset was split into batches of 5,000. A population specific reference panel consisting of 2,695 WGS samples was utilized for imputation and standard Beagle hg38 recombination maps were used. Based on principal component analysis, samples who were not of European ancestry were removed. Duplicate and monozygous twin detection was performed with KING 2.2.7^50^, and one sample was removed out of the pair of duplicates.

Association analysis in EstBB was carried out for all variants with an INFO score > 0.4 using the additive model as implemented in REGENIE v3.0.3 with standard binary trait settings^46^. Logistic regression was carried out with adjustment for current age, age^2^, sex and 10 PCs as covariates, analyzing only variants with a minimum minor allele count of 2.

### FinnGen

Finnish ancestry samples from Finnish public-private research project FinnGen were used^51^. FinnGen release 12 comprises genome information with digital healthcare data on ∼10% of Finnish population (https://www.finngen.fi/en). Individuals in the FinnGen release 12 with the International Classification of Diseases Tenth Revision (ICD-10) diagnosis code U07* for SARS-CoV-2 infection (U07.1 or U07.2, virus identified or not identified, respectively) were defined as SARS-CoV-2-infected. For the GWAS of Omicron, individuals were grouped by the diagnosis date of their first SARS-CoV-2 infection. As Omicron variants became the main lineage in December 2021 in Finland, we defined individuals with their first SARS-CoV-2 diagnosis date starting from 1.1.2022 as Omicron cases (N = 61,393). Individuals with no SARS-CoV-2 diagnosis were used as controls (N = 399,149). For the comparison with earlier SARS-CoV-2 variants, individuals with diagnosis dates before or in November 2021 were defined as controls (N = 35,594).

Diagnosis dates in FinnGen data are pseudonymised by +-2 weeks, thus individuals with their first SARS-CoV-2 diagnosis during the Delta-Omicron transition period, December 2021, were excluded from the earlier SARS-CoV-2 controls.

FinnGen samples were genotyped with ThermoFisher, Illumina and Affymetrix arrays. Imputation was performed using the Finnish population-specific imputation panel SISu v4 (version 4.2). FinnGen data (180,000 SNPs) was compared to 1k Genome Project data, with a Bayesian algorithm detecting PCA outliers. 35,371 samples were detected as either non-Finnish ancestry or twins/duplicates with relations to other samples and thus excluded. Of the 500,737 non-duplicate population inlier samples from PCA, 355 samples were excluded from analysis because of missing minimum phenotype data, and 34 samples because of failing sex check with F thresholds of 0.4 and 0.7. A total of 500,348 samples (282,064 [56.4%] females and 218,284 [43.6%] males) were accepted for phenotyping for the GWAS analyses.

Case vs control GWAS analyses were performed using REGENIE (version 2.2.4)^46^. Logistic regression was adjusted for age (at death or end of registry follow-up), sex, the first 10 principal components, and genotyping batches. Firth approximation test was applied for variants with an initial p-value of less than 0.01 and standard error was computed based on effect size and likelihood ratio test p-value (regenie options --firth --approx --pThresh 0.01 --firth-se).

### Mass General Brigham Biobank

The Mass General Brigham (MGB) Biobank, formerly known as the Partners Biobank, is a hospital-based cohort study produced by the MGB healthcare network located in Boston, MA. The MGB Biobank contains data from patients in multiple primary care facilities, as well as tertiary care centers located in the greater Boston area. Participants of the study are recruited from inpatient stays, emergency department environments, outpatient visits, and through a secure online portal available to patients. Recruitment and consent are fully translatable to Spanish in order to promote a greater patient diversity. This allows for a systematic enrollment of diverse patient groups which is reflective of the population receiving care through the MGB network. Recruitment for the biobank began in 2009 and is still actively recruiting. The recruitment strategy has been described previously^52^.

Cases for SARS-CoV-2 infection with Omicron variants were ascertained from the Mass General Brigham Biobank (data access April 23, 2024). Individuals with a SARS-CoV-2 infection were curated by the biobank and represent those who presented to the hospital system with a positive infection control flag, presumed infection control flag, and/or a SARS-CoV-2 RNA positive test result. Cases of Omicron infections were defined as individuals presenting with a SARS-CoV-2 infection after January 1, 2022. Control definition included individuals in the MGB Biobank without any report of infection. For the comparison with infections with earlier variants, controls were defined as individuals with a SARS-CoV-2 infection prior to December 1, 2021 only.

The MGB Biobank genotyped 53,297 participants on the Illumina Global Screening Array (‘GSA’) and 11,864 on Illumina Multi-Ethnic Global Array (“MEG”). The GSA arrays captured approximately 652K SNPs and short indels, while the MEG arrays captured approximately 1.38M SNPs and short indels. These genotypes were filtered for high missingness (>2%) and variants out of HWE (P < 1e-12), as well as variants with an AF discordant (P<1e-150) from a synthesized AF calculated from GnomAD subpopulation frequencies and a genomewide GnomAD model fit of the entire cohort. This resulted in approximately 620K variants for GSA and 1.15M for MEG. The two sets of genotypes were then separately phased and imputed on the TOPMed imputation server (Minimac4 algorithm) using the TOPMed r2 reference panel. The resultant imputation sets were both filtered at an R2 > 0.4 and a MAF > .001, and then the two sets were merged/intersected resulting in approximately 19.5M GRCh38 autosomal variants. The sample set for analysis here was then restricted to just those classified as EUR according to a random-forest classifier trained with the Human Genome Diversity Project (HGDP) as the reference panel, with the minimum probability for assignment to an ancestral group of 0.5, in 19/20 iterations of the model^52^. To correct for population stratification, PCs were computed in genetically European participants. Association analysis was performed with variants using REGENIE (v3.2.8) with adjustment for age, age squared, sex, chip, tranche, and PC 1-10.

### Meta-analysis

Initial REGENIE results were filtered based on a MAF > 0.1% and an INFO score > 0.8 and analyzed in METAL (version 2011.03.25)^53^ by the inverse-variance method with genomic control applied to the input files. The results from the meta-analysis were filtered for SNPs present in all three major cohorts, resulting in a total of 8,669,333 SNPs, of which 436,360 did not have results for the MGB cohort (including all 224,900 SNPs from chromosome X).

### Linkage disequilibrium (LD) calculations

When not otherwise stated, LD between SNPs was calculated in LDpair (https://ldlink.nih.gov/?tab=ldpair) based on the 5 European ancestry groups from Utah, Italy, Finland, Great Britain, and Spain. In cases were one of the SNPs was not available in the 1000 genome reference panel, LD was calculated based on the Danish study cohort.

### STRING functional protein association network analysis

We obtained functional protein association networks from STRING database v12^19^, which we visualized in Cytoscape v3.10^54^ using stringApp v2.1.1^55^. Initially, we selected one gene per locus, based on physical proximity to the lead SNP and additional evidence from FUMA results and used the default confidence score threshold of 0.4 (https://version-12-0.string-db.org/cgi/network?networkId=bnOf0kS7q9qc).

One functionality of STRING is expanding a given network with a user-defined number of interactors at a specific degree of selectivity. We expanded the initial network with 15 interactors, setting the selectivity parameter to the maximum value of 1, allowing us to identify proteins that primarily interact with the current network, and are not hubs of the entire STRING network. The genes for some of the 15 retrieved interactors were located at the same locus, or at a locus already represented in the initial network. In these cases, we selected only the entry with most interactions in the network and removed the other proteins at this locus from the network.

### Ethics

The Copenhagen Hospital Biobank provides biological left-over samples from routine blood analyses and the patients were not asked for informed consent before inclusion. Instead, patients were informed about the opt-out possibility to have their biological specimens excluded from use in research. Individuals from the exclusion register (Vævsanvendelsesregistret) were excluded from the study. For the Danish Blood Donor Study, informed consent was obtained from all participants. Both studies are part of a COVID-19 protocol approved by the National Ethics Committee (H-21030945) and the Danish Data Protection Agency (P-2020-356).

EFTER-COVID was conducted as a surveillance study as part of Statens Serum Institut’s advisory tasks for the Danish Ministry of Health. According to Danish law, these national surveillance activities do not require approval from an ethics committee. Participation in the study was voluntary and the invitation letter contained information about participants’ rights under the Danish General Data Protection Regulation (rights to access data, rectification, deletion, restriction of processing and objection). After reading this information, it was considered informed consent when participants read the information and agreed, and then continued to fill in the questionnaires.

The activities of the Estonian Biobank (EstBB) are regulated by the Human Genes Research Act, which was adopted in 2000 specifically for the operations of EstBB. Individual level analysis with EstBB data was carried out under ethical approval 1.1-12/624 from the Estonian Committee on Bioethics and Human Research (Estonian Ministry of Social Affairs), using data according to release application 6-7/GI/5933 from the Estonian Biobank.

Study subjects in FinnGen provided informed consent for biobank research, based on the Finnish Biobank Act. Alternatively, separate research cohorts, collected prior the Finnish Biobank Act came into effect (in September 2013) and start of FinnGen (August 2017), were collected based on study-specific consents and later transferred to the Finnish biobanks after approval by Fimea (Finnish Medicines Agency), the National Supervisory Authority for Welfare and Health. Recruitment protocols followed the biobank protocols approved by Fimea. The Coordinating Ethics Committee of the Hospital District of Helsinki and Uusimaa (HUS) statement number for the FinnGen study is Nr HUS/990/2017. The FinnGen study is approved by the Finnish Institute for Health and Welfare and other authorities (a complete overview of permissions is given in the **Supplementary Note**).

The Mass General Brigham (MGB) Biobank, formerly known as the Partners Biobank, is a hospital-based cohort study produced by the MGB healthcare network located in Boston, MA. The MGB Biobank contains data from patients in multiple primary care facilities, as well as tertiary care centers located in the greater Boston area. Participants of the study are recruited from inpatient stays, emergency department environments, outpatient visits, and through a secure online portal available to patients. Recruitment and consent are fully translatable to Spanish in order to promote a greater patient diversity. This allows for a systematic enrollment of diverse patient groups which is reflective of the population receiving care through the MGB network. Recruitment for the biobank began in 2009 and is still actively recruiting. The recruitment strategy has been described previously^52^. For the MGB Biobank, all patients provide written consent upon enrollment. Furthermore, the MGB cohort included test verified SARS-CoV-2 infection data with time of diagnosis. The present study protocol was approved by the MGB Institutional Review Board (#2018P002276).

## Data Availability

GWAS meta-analysis summary statistics are available for interactive plotting and viewing via LocusZoom56 (https://my.locuszoom.org/gwas/962995/?token=0fa64e3ae9d445ddb7b5bd3a32c6b4d6) and will be made public and deposited at https://www.danishnationalbiobank.com/gwas/ after publication.
Complete FUMA results (including the MAGMA analysis) are available online (https://fuma.ctglab.nl/browse/475677).

https://fuma.ctglab.nl/browse/475677

## Acknowledgments

We thank all participants and staff related to the Copenhagen Hospital Biobank, Danish Blood Donor Study, EFTER-COVID, FinnGen, Estonian Biobank and Mass General Brigham Biobank for their contribution to this research.

This work was supported by research grants from Sygeforsikringen “danmark” (2020-0178) and the EU Horizon REACT study (101057129). The Copenhagen Hospital Biobank (CHB) was funded by grants from Novo Nordisk Foundation (NNF23OC0082015) and Rigshospitalet Research Council (Framework grant) and by Novo Nordisk Foundation CHALLENGE grant (NNF17OC0027594). The Danish Blood Donor Study (DBDS) was funded by the Danish Council for Independent Research - Medical Sciences and the Danish Administrative Regions (Bio- and Genome Bank Denmark). S. Bliddal received grants from the Novo Nordisk Foundation (NNF22OC0077221,NNF23OC0087269), L.A.N.C. was funded by NordForsk (project number 105668 and 138929), B.F. was partially supported by the Novo Nordisk Foundation (NNF17OC0027594).

The Danish Departments of Clinical Microbiology (KMA) and Statens Serum Institut carried out laboratory analyses, registration, and release of the national SARS-CoV-2 surveillance data for the present study.

The work of the Estonian Genome Center, University of Tartu, was funded by the European Union through Horizon 2020 research and innovation program under grants no. 894987, 101137201 and 101137154, and Estonian Research Council Grant PRG1291. The Estonian Genome Center analyses were partially carried out in the High Performance Computing Center, University of Tartu.

We want to acknowledge the participants and investigators of the FinnGen study. The FinnGen project is funded by two grants from Business Finland (HUS 4685/31/2016 and UH 4386/31/2016) and the following industry partners: AbbVie Inc., AstraZeneca UK Ltd, Biogen MA Inc., Bristol Myers Squibb (and Celgene Corporation & Celgene International II Sàrl), Genentech Inc., Merck Sharp & Dohme LCC, Pfizer Inc., GlaxoSmithKline Intellectual Property Development Ltd., Sanofi US Services Inc., Maze Therapeutics Inc., Janssen Biotech Inc, Novartis AG, and Boehringer Ingelheim International GmbH. Following biobanks are acknowledged for delivering biobank samples to FinnGen: Auria Biobank (www.auria.fi/biopankki), THL Biobank (www.thl.fi/biobank), Helsinki Biobank (www.helsinginbiopankki.fi), Biobank Borealis of Northern Finland (https://www.ppshp.fi/Tutkimus-ja-opetus/Biopankki/Pages/Biobank-Borealis-briefly-in-English.aspx), Finnish Clinical Biobank Tampere (www.tays.fi/en-US/Research_and_development/Finnish_Clinical_Biobank_Tampere), Biobank of Eastern Finland (www.ita-suomenbiopankki.fi/en), Central Finland Biobank (www.ksshp.fi/fi-FI/Potilaalle/Biopankki), Finnish Red Cross Blood Service Biobank (www.veripalvelu.fi/verenluovutus/biopankkitoiminta), Terveystalo Biobank (www.terveystalo.com/fi/Yritystietoa/Terveystalo-Biopankki/Biopankki/) and Arctic Biobank (https://www.oulu.fi/en/university/faculties-and-units/faculty-medicine/northern-finland-birth-cohorts-and-arctic-biobank). All Finnish Biobanks are members of BBMRI.fi infrastructure (https://www.bbmri-eric.eu/national-nodes/finland/). Finnish Biobank Cooperative -FINBB (https://finbb.fi/) is the coordinator of BBMRI-ERIC operations in Finland. The Finnish biobank data can be accessed through the Fingenious® services (https://site.fingenious.fi/en/) managed by FINBB.

We thank the Mass General Brigham Biobank for providing samples, genomic data, and health information data for genetic analyses. We acknowledge support from NIH R01AI170850 (J.V.), NIH R35GM146839 (J.M.L.) and NIH R01HG012810 (J.M.L.).

## Data availability

GWAS meta-analysis summary statistics are available for interactive plotting and viewing via LocusZoom^56^ (https://my.locuszoom.org/gwas/962995/?token=0fa64e3ae9d445ddb7b5bd3a32c6b4d6) and will be made public and deposited at https://www.danishnationalbiobank.com/gwas/ after publication. Complete FUMA results (including the MAGMA analysis) are available online (https://fuma.ctglab.nl/browse/475677).

## Code availability

Code for the analysis will be available under OmicronCode at our GitHub site (https://github.com/FeenstraLab).

## Tables and Figures

**Table 1** Lead variants for the 13 genome-wide significant loci in the GWAS of Omicron infection vs. no known infection.

**Figure 1** Manhattan plot for GWAS of Omicron infection vs. no known infection.

**Figure 2** STRING Network

## Supplementary Tables

**Supplementary Table 1** Results from GWAS of Omicron infection vs. no known infection for 51 SNPs reported by the HGI as associated with SARS-CoV-2 infection and/or hospitalization.

**Supplementary Table 2** Results for the lead SNPs from Table 1 in a GWAS of cases of SARS-CoV-2 infection with the Omicron variant vs. controls with a SARS-CoV-2 infection with earlier variants.

**Supplementary Table 3** Results for the lead SNPs from Table 1 in a GWAS of breakthrough infections.

**Supplementary Table 4** Results for the lead SNPs from Table 1 in a GWAS of influenza.

**Supplementary Table 5** PheWAS results for the lead SNPs from Table 1 for 2,470 phenotypes provided in FinnGen release 12.

**Supplementary Table 6** Reactome gene-sets with an adjusted P < 0.05 in the MAGMA analysis.

**Supplementary Table 7** Genetic correlations of our GWAS with GWAS for 1,461 traits implemented in the Complex Traits Genetics Virtual Lab.

**Supplementary Table 8** Basic descriptive statistics on age and sex of cases and controls from all cohorts.

## Supplementary Information

**Supplementary Figure 1** Forest plots for four SNPs with signs of heterogeneity of effect.

**Supplementary Figure 2** Distribution of Omicron cases in the four GWAS cohorts.

**Supplementary Figure 3** Distribution of SARS-CoV-2 variants from December 2021 to May 2023 in the four countries of the study cohorts.

## Supplementary Notse

List of members of the DBDS Genomic Consortium, Estonian Biobank Research Team, and FinnGen, permissions for FinnGen.

## References

1. COVID-19 cases | WHO COVID-19 dashboard. https://data.who.int/dashboards/covid19/cases (accession date: 14 November 2024).

2. Kanai, M. et al. A second update on mapping the human genetic architecture of COVID-19. Nat. 2023 6217977 621, E7–E26 (2023).

3. Markov, P. V. et al. The evolution of SARS-CoV-2. Nat. Rev. Microbiol. 2023 216 21, 361–379 (2023).

4. Azad, M. B., Wade, K. H. & Timpson, N. J. FUT2 secretor genotype and susceptibility to infections and chronic conditions in the ALSPAC cohort. Wellcome Open Res. 3, (2018).

5. Varki, A. Sialic acids in human health and disease. Trends Mol. Med. 14, 351–360 (2008).

6. Kosmicki, J. A. et al. Genetic risk factors for COVID-19 and influenza are largely distinct. Nat. Genet. 56, 1592–1596 (2024).

7. Emilsson, V. et al. Co-regulatory networks of human serum proteins link genetics to disease. Science 361, (2018).

8. Keaton, J. M. et al. Genome-wide analysis in over 1 million individuals of European ancestry yields improved polygenic risk scores for blood pressure traits. Nat. Genet. 56, 778–791 (2024).

9. Sakaue, S. et al. A cross-population atlas of genetic associations for 220 human phenotypes. Nat. Genet. 53, 1415–1424 (2021).

10. Shrine, N. et al. New genetic signals for lung function highlight pathways and chronic obstructive pulmonary disease associations across multiple ancestries. Nat. Genet. 51, 481–493 (2019).

11. Paganini, J. et al. HLA-F transcriptional and protein differential expression according to its genetic polymorphisms. HLA 102, 578–589 (2023).

12. Lin, A. & Yan, W. H. The emerging roles of human leukocyte antigen-F in immune modulation and viral infection. Front. Immunol. 10, 449250 (2019).

13. Noh, H. E. & Rha, M. S. Mucosal Immunity against SARS-CoV-2 in the Respiratory Tract. Pathog. 2024, Vol. 13, Page 113 13, 113 (2024).

14. Gram, M. A. et al. Patterns of testing in the extensive Danish national SARS-CoV-2 test set-up. PLoS One 18, (2023).

15. Alcalde-Herraiz, M. et al. Genome-wide association studies of COVID-19 vaccine seroconversion and breakthrough outcomes in UK Biobank. Nat. Commun. 2024 151 15, 1–10 (2024).

16. Watanabe, K., Taskesen, E., van Bochoven, A. & Posthuma, D. Functional mapping and annotation of genetic associations with FUMA. Nat. Commun. 8, 1826 (2017).

17. Wang, G., Sarkar, A., Carbonetto, P. & Stephens, M. A Simple New Approach to Variable Selection in Regression, with Application to Genetic Fine Mapping. J. R. Stat. Soc. Ser. B Stat. Methodol. 82, 1273–1300 (2020).

18. de Leeuw, C. A., Mooij, J. M., Heskes, T. & Posthuma, D. MAGMA: generalized gene-set analysis of GWAS data. PLoS Comput. Biol. 11, e1004219–e1004219 (2015).

19. Szklarczyk, D. et al. The STRING database in 2023: protein-protein association networks and functional enrichment analyses for any sequenced genome of interest. Nucleic Acids Res. 51, D638– D646 (2023).

20. Milacic, M. et al. The Reactome Pathway Knowledgebase 2024. Nucleic Acids Res. 52, D672–D678 (2024).

21. Kanehisa, M., Goto, S., Sato, Y., Furumichi, M. & Tanabe, M. KEGG for integration and interpretation of large-scale molecular data sets. Nucleic Acids Res. 40, (2012).

22. Fernández-Ponce, C. et al. The Role of Glycosyltransferases in Colorectal Cancer. Int. J. Mol. Sci. 22, (2021).

23. Zhu, J., Dingess, K. A., Mank, M., Stahl, B. & Heck, A. J. R. Personalized Profiling Reveals Donor-and Lactation-Specific Trends in the Human Milk Proteome and Peptidome. J. Nutr. 151, 826–839 (2021).

24. Matrosovich, M., Matrosovich, T., Carr, J., Roberts, N. A. & Klenk, H.-D. Overexpression of the α-2,6-Sialyltransferase in MDCK Cells Increases Influenza Virus Sensitivity to Neuraminidase Inhibitors. J. Virol. 77, 8418 (2003).

25. Kim, C. H. SARS-CoV-2 Evolutionary Adaptation toward Host Entry and Recognition of Receptor O-Acetyl Sialylation in Virus–Host Interaction. Int. J. Mol. Sci. 2020, Vol. 21, Page 4549 21, 4549 (2020).

26. Sun, X. L. The role of cell surface sialic acids for SARS-CoV-2 infection. Glycobiology 31, 1245 (2021).

27. Mykytyn, A. Z. et al. SARS-CoV-2 Omicron entry is type II transmembrane serine protease-mediated in human airway and intestinal organoid models. J. Virol. 97, (2023).

28. Willett, B. J. et al. SARS-CoV-2 Omicron is an immune escape variant with an altered cell entry pathway. Nat. Microbiol. 2022 78 7, 1161–1179 (2022).

29. Laine, L., Skön, M., Väisänen, E., Julkunen, I. & Österlund, P. SARS-CoV-2 variants Alpha, Beta, Delta and Omicron show a slower host cell interferon response compared to an early pandemic variant. Front. Immunol. 13, 1016108 (2022).

30. Flagg, M. et al. Low level of tonic interferon signalling is associated with enhanced susceptibility to SARS-CoV-2 variants of concern in human lung organoids. Emerg. Microbes Infect. 12, (2023).

31. Halfmann, P. J. et al. SARS-CoV-2 Omicron virus causes attenuated disease in mice and hamsters. Nat. 2022 6037902 603, 687–692 (2022).

32. Armando, F. et al. SARS-CoV-2 Omicron variant causes mild pathology in the upper and lower respiratory tract of hamsters. Nat. Commun. 2022 131 13, 1–15 (2022).

33. Watanabe, Y., Bowden, T. A., Wilson, I. A. & Crispin, M. Exploitation of glycosylation in enveloped virus pathobiology. Biochim. Biophys. acta. Gen. Subj. 1863, 1480–1497 (2019).

34. Casalino, L. et al. Beyond shielding: The roles of glycans in the SARS-CoV-2 spike protein. ACS Cent. Sci. 6, 1722–1734 (2020).

35. Shajahan, A., Pepi, L. E., Kumar, B., Murray, N. B. & Azadi, P. Site specific N- and O-glycosylation mapping of the spike proteins of SARS-CoV-2 variants of concern. Sci. Rep. 13, 10053 (2023).

36. Wang, D. et al. Enhanced Surface Accessibility of SARS-CoV-2 Omicron Spike Protein Due to an Altered Glycosylation Profile. ACS Infect. Dis. 13, 23 (2024).

37. Xie, Y. & Butler, M. Quantitative profiling of N-glycosylation of SARS-CoV-2 spike protein variants. Glycobiology 33, 188–202 (2023).

38. Huang, C. et al. The effect of N-glycosylation of SARS-CoV-2 spike protein on the virus interaction with the host cell ACE2 receptor. iScience 24, (2021).

39. Zheng, L. et al. Characterization and Function of Glycans on the Spike Proteins of SARS-CoV-2 Variants of Concern. Microbiol. Spectr. 10, (2022).

40. Wang, S. et al. Sequential glycosylations at the multibasic cleavage site of SARS-CoV-2 spike protein regulate viral activity. Nat. Commun. 2024 151 15, 1–17 (2024).

41. Zhang, L. et al. Furin cleavage of the SARS-CoV-2 spike is modulated by O-glycosylation. Proc. Natl. Acad. Sci. U. S. A. 118, e2109905118 (2021).

42. Gonzalez-Rodriguez, E. et al. O-Linked Sialoglycans Modulate the Proteolysis of SARS-CoV-2 Spike and Likely Contribute to the Mutational Trajectory in Variants of Concern. ACS Cent. Sci. 9, 393–404 (2023).

43. Sørensen, A. I. V. et al. Cohort profile: EFTER-COVID - a Danish nationwide cohort for assessing the long-term health effects of the COVID-19 pandemic. BMJ Open 14, (2024).

44. Sørensen, E. et al. Data Resource Profile: The Copenhagen Hospital Biobank (CHB). Int. J. Epidemiol. 50, 719–720e (2021).

45. Hansen, T. F. et al. DBDS Genomic Cohort, a prospective and comprehensive resource for integrative and temporal analysis of genetic, environmental and lifestyle factors affecting health of blood donors. BMJ Open 9, e028401 (2019).

46. Mbatchou, J. et al. Computationally efficient whole-genome regression for quantitative and binary traits. Nat. Genet. 2021 537 53, 1097–1103 (2021).

47. Leitsalu, L. et al. Cohort Profile: Estonian Biobank of the Estonian Genome Center, University of Tartu. Int. J. Epidemiol. 44, 1137–1147 (2015).

48. Mitt, M. et al. Improved imputation accuracy of rare and low-frequency variants using population-specific high-coverage WGS-based imputation reference panel. Eur. J. Hum. Genet. 25, 869–876 (2017).

49. Browning, B. L., Tian, X., Zhou, Y. & Browning, S. R. Fast two-stage phasing of large-scale sequence data. Am. J. Hum. Genet. 108, 1880–1890 (2021).

50. Manichaikul, A. et al. Robust relationship inference in genome-wide association studies. Bioinformatics 26, 2867–2873 (2010).

51. Kurki, M. I. et al. FinnGen provides genetic insights from a well-phenotyped isolated population. Nat. 2023 6137944 613, 508–518 (2023).

52. Dashti, H. S. et al. Interaction of obesity polygenic score with lifestyle risk factors in an electronic health record biobank. BMC Med. 20, (2022).

53. Willer, C. J., Li, Y. & Abecasis, G. R. METAL: Fast and efficient meta-analysis of genomewide association scans. Bioinformatics (2010) doi:10.1093/bioinformatics/btq340.

54. Shannon, P. et al. Cytoscape: a software environment for integrated models of biomolecular interaction networks. Genome Res. 13, 2498–2504 (2003).

55. Doncheva, N. T. et al. Cytoscape stringApp 2.0: Analysis and Visualization of Heterogeneous Biological Networks. J. Proteome Res. 22, 637–646 (2023).

56. Boughton, A. P. et al. LocusZoom.js: interactive and embeddable visualization of genetic association study results. Bioinformatics 37, 3017–3018 (2021).

